# A deep learning approach with event-related spectral EEG data in attentional deficit hyperactivity disorder

**DOI:** 10.1101/19005611

**Authors:** Laura Dubreuil-Vall, Giulio Ruffini, Joan A. Camprodon

## Abstract

Attention deficit hyperactivity disorder (ADHD) is a heterogeneous neurodevelopmental disorder that affects 5% of the pediatric and adult population worldwide. The diagnosis remains essentially clinical, based on history and exam, with no available biomarkers. In this paper, we describe a deep convolutional neural network (DCNN) for ADHD classification derived from the time-frequency decomposition of electroencephalography data (EEG), particularly of event-related potentials (ERP) during the Flanker Task collected from 20 ADHD adult patients and 20 healthy controls (HC). The model reaches a classification accuracy of 88%, superior to resting state EEG spectrograms and with the key advantage, compared with other machine learning approaches, of avoiding the need for manual selection of EEG spectral or channel features. Finally, through the use of feature visualization techniques, we show that the main features exciting the DCNN nodes are a decreased power in the alpha band and an increased power in the delta-theta band around 100ms for ADHD patients compared to HC, suggestive of attentional and inhibition deficits, which have been previously suggested as pathophyisiological signatures of ADHD. While confirmation with larger clinical samples is necessary, these results highlight the potential of this methodology to develop CNS biomarkers of practical clinical utility.

## Introduction

Attention-deficit hyperactivity disorder (ADHD) is a neurodevelopmental disorder characterized by deficits in attention, impulsivity (motor and non-motor) and executive dysfunction. It is associated with high morbidity and disability^1,2^, and affects up to 5% of adults worldwide^3-5^. The diagnosis of ADHD remains essentially clinical, based on history and exam. It can be supported by neuropsychological assessments, but given the heterogeneous cognitive profiles in patients with ADHD, these provide a supportive, not fully diagnostic, function. Significantly, there are many different conditions that present with disordered attention, impulsivity and dysexecutive syndromes, and the range of normal cognitive profiles with variable strengths and weaknesses in these domains is wide, often complicating the differential diagnosis. Hence, a biomarker to reduce the inherent uncertainty of clinical diagnosis would be of great value.

Electroencephalographic (EEG) signals contain rich information associated with functional dynamics in the brain. The use of EEG in ADHD began more than 75 years ago with Jasper et al.^6^ reporting an increase in the EEG power of low frequencies in fronto-central areas. Since then, human electrophysiological studies using EEG spectral analyses and event-related potentials (ERPs) have established relevant signatures of executive dysfunction in ADHD^7^. In contrast to spontaneous EEG, ERPs reflect changes in the electrical activity of the brain that are time-locked to the occurrence of a specific event, that is, a response to a discrete external stimulus or an internal mental process^8^. ERPs also provide non-invasive neurophysiological measurements with high temporal resolution, allowing to assess dysfunctional brain dynamics, including cognitive processes that may not be apparent at the behavioral level^9,10^. Indeed, ERPs are commonly used clinically in neurophysiological diagnostic units to support the assessment of neuropsychiatric disorders (e.g., multiple sclerosis^11^) and sensory disorders (e.g., screening of neonates for hearing impairments^12^).

Artificial neural networks (ANNs) have recently become a promising application of artificial intelligence (AI) in healthcare^13^. Machine learning, a subtype of AI, and deep learning, a specialized sub-field of machine learning, have been increasingly used in clinical research with promising results. Machine learning can be described as the practice of using algorithms to train a system by using large amounts of data, with the goal of giving it the ability to learn how to perform a specific task, and then make an accurate classification or prediction. Deep learning is a subset of machine learning algorithms that break down the tasks in smaller units (neural networks, NNs) often providing higher levels of accuracy.

NNs are characterized by their network architecture, defined by the anatomical layout of its connected processing units, the artificial “neurons”, according to a loss or optimization function that specifies the overall goal of the learning process. Connections are “trained”, or taught how to do the desired task, by using a training algorithm that iteratively changes parameters of the NN such that the target function is ultimately optimized based on the inputs the NN receives. There are different types of NNs with different designs and architectures derived from different principles, or conceived for different purposes. The most basic ones are the feed-forward NNs (FNNs), in which activity is propagated unidirectionally layer-by-layer from the input up to the output stage, with no feedback connections within or between layers. We have previously used a specific type of FFNs (feed-forward autoencoders) for the analysis of EEG data with promising outcomes^14^. Recurrent Neural Networks (RNN) are another type of NN that, unlike FFNs, are based on architectures with feedback (“recurrent”) connections within or between layers. In related work, we used Echo State Networks (ESNs), a particular type of RNN, to classify Parkinson patients from HC using EEG time-frequency decompositions^15^ with successful results. The main limitation of RNNs is, however, their computational cost^16^. In addition, one of the main critics to deep NN is their “black-box” nature, i.e., the difficulty in tracing a prediction back to which features are important and understanding how the network reached the final output, which will be later addressed in this study.

Previous studies have successfully classified ADHD patients from HC using machine learning techniques with accuracies of more than 90%^17-23^, but the selection of disease-characterizing features from EEG was done manually after an extensive search in the frequency or time domain. However, EEG signals exhibit non-linear dynamics (chaotic signals that do not behave linearly and cannot be represented as combination of basic sub-signals) and non-stationarity across temporal scales (signals with a mean and variance that do not stay constant over time) that cannot be studied properly using classical machine learning approaches. There is a need for tools capable of capturing the rich spatiotemporal hierarchical structures hidden in these signals. In a previous study^24^, we trained a machine learning system with pre-defined complexity metrics of time-frequency decompositions of EEG data that showed statistically significant differences between REM Sleep Behavior Disorder (RBD) patients and HC, indicating that such metrics may be useful for classification or scoring. While this approach is useful in several domains, it would be advantageous to use methods where the relevant features are found directly by the algorithms instead of pre-defining them manually.

With the goal of building a discrimination system that can classify ADHD patients from HCs, here we explore a deep learning approach inspired by recent successes in image classification using Deep Convolutional Neural Networks (DCNNs), a particular type of NN designed to exploit compositional and translationally invariant features in the data that are present in EEG, i.e., features that are recognizable even if their appearance varies in some way^16^. These networks were originally developed to deal with image data (2D arrays) from different channels or audio data^25^, and more recently, EEG data^26,27^. Similarly, here we train a DCNN with multi-channel two-dimensional time-frequency maps (spectrograms or 2D time-frequency maps), representing EEG spectral dynamics as images with the equivalent image depth provided by multiple EEG channels. These networks treat EEG-channel data as an audio file, and our approach mimics similar uses of deep networks in that domain. Specifically, we use a similar strategy as the one presented by Ruffini et al.^28^, but instead of using spontaneous EEG spectrograms, we use ERP spectrograms (also called Event-Related Spectral perturbation, ERSP) recorded during a Flanker-Eriksen Task (EFT), a well-established experimental task to assess sustained attention, conflict monitoring and response inhibition. Our assumption is that relevant qualities of ERP data are contained in compositional features embedded in this time-frequency representation. Particularly, we expect that DCNNs may be able to efficiently learn to identify features in the time-frequency domain associated to event-related bursting across frequency bands that may help separate classes, similar to what is known as “bump analysis”^29^. For comparison purposes, we also trained a RNN based on Long Short-Term Memory (LSTM) networks, which can learn long sequences of data but require higher computational demands, and a Shallow Neural Network (SNN) as a baseline, a more basic type of network with only one layer. We also compared the performance of the ERSP data with a dataset of spontaneous EEG data recorded while the participants were at resting state. Lastly, we propose the utilization of deep learning visualization techniques for the mechanistic interpretation of results, particularly the method popularly known as DeepDream^30^. This is important to identify pathophysiological features driving the translational and clinical value of the application, and for the optimized further development and acceptance of such techniques in the clinical domain, where black-box approaches have been extensively criticized.

## Methods

### Participants

A total of 40 participants including 20 healthy adults (10 males, 10 females) and 20 ADHD adult patients (10 males, 10 females) participated in the present study (Table 1). The inclusion criteria for ADHD patients consisted of a diagnosis of ADHD made by a board-certified clinician according to the Diagnostic and Statistical Manual of Mental Disorders, Fifth Edition (DSM-5)^31^. Symptom profiles and severity were assessed with the Adult ADHD Self-Report Scale (ASRS-v1.1)^32^. Patients were either off stimulant medications or, if undergoing treatment with stimulants, were asked to discontinue two days prior to the experiment, under a physician-guided protocol, and allowed to resume afterwards. Psychiatric comorbidities were allowed as long as ADHD was the primary diagnosis. Psychosis, bipolar disorder, substance use disorder and neurological conditions were exclusion criteria. Healthy participants were included if they did not have any psychiatric or neurologic condition and were not taking any psychoactive medications. All participants gave informed and written consent for participation. The study was approved by the Partners HealthCare System’s Institutional Review Board and all experiments were performed in accordance with relevant guidelines and regulations at Massachusetts General Hospital.

**Table 1.** Participant characteristics

|  | ADHD (n=20) | HC (n=20) | Significance |
| --- | --- | --- | --- |
| Demographic | mean (SD)* | mean (SD)* | p value (T test) |
| Age | 43.85 (14.78) | 29.90 (10.77) | 0.0006 |
| Females | 10 (50%) | 10 (50%) | 0.5 |
| **Baseline Scores** |  |  |  |
| ASRS | 62.6 (9.17) | 36.47 (11.33) | <0.0001 |
| **Current medications – N (%)** |  |  |  |
| No medication | 11 (55%) |  |  |
| Adderall | 2 (15%) |  |  |
| Vyvanse | 2 (10%) |  |  |
| Concerta | 1 (5%) |  |  |
| Verapamil | 1 (5%) |  |  |
| Aspirin | 1 (5%) |  |  |
| Levothyroxine | 1 (5%) |  |  |
| Modafinil | 1 (5%) |  |  |

Abbreviations. SD: Standard Deviation
(\*) All figures are mean (Standard Deviation) unless otherwise specified.

### Experimental Task: Eriksen-Flanker task (EFT)

Each patient underwent three identical experimental sessions separated by 1-2 weeks in which they performed the Eriksen-Flanker task (EFT) (Figure 1) while EEG data was recorded. The EFT is a classic behavioral paradigm in which subjects must attend and respond to the direction of a central arrow that is surrounded (“flanked”) by distracting stimuli. The flanking arrows can either have the same (congruent trials) or opposing (incongruent trials) orientation as the central one. Participants are instructed to press the left or right arrow buttons in a keyboard following the direction of the central arrow, ignoring the flankers. In this study there were a total of 140 trials, and each subject had a different, fully random sequence of congruent and incongruent trials, with 2 congruent trials for each incongruent trial, in order to build a tendency towards the prepotent congruent responses and thus increase the difficulty of conflict detection in incongruent trials. Only incongruent trials were used for classification purposes, as they are the ones that most elicit the conflict-related ERP components that characterize the executive function subtasks of selective attention, inhibition and cognitive control^33^, primarily impaired in ADHD. The accuracy (percentage of correct/incorrect responses) and the reaction time (RT) were measured for each trial, while also recording EEG data during the task. RT of single trials was introduced into a Generalized Linear Model with Mixed Effects (GLMM) with a Gamma distribution, with Group as a fixed factor (ADHD/HC) and Subject ID as a random intercept. Accuracy was also modeled using a generalized logistic regression model with mixed effects and a binomial distribution.

**Figure 1.**
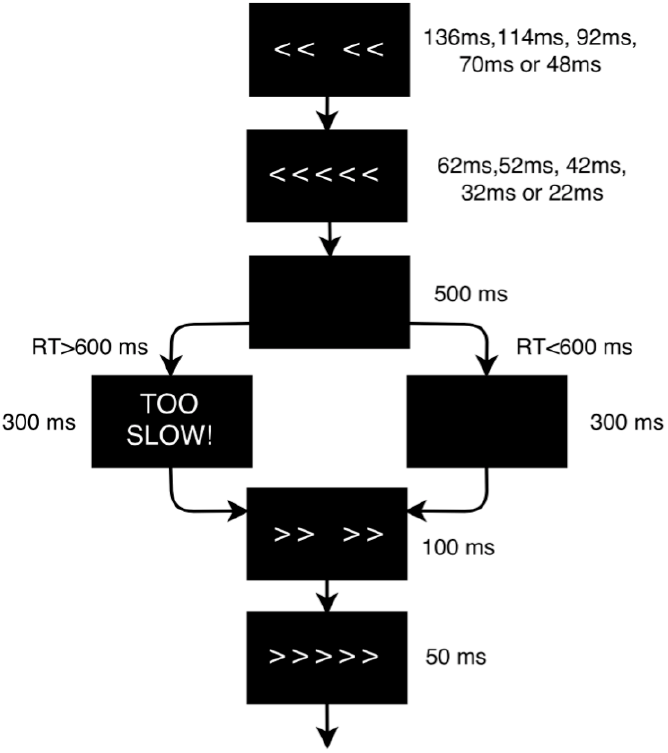
Flanker task design scheme. The flanker arrows were first presented alone for duration of 136ms, 114ms, 92ms, 70ms or 48ms depending on the baseline performance of each subject, and were then joined by the target arrow for 62ms, 52ms, 42ms, 32ms or 22ms, respectively (the duration of the stimuli was adjusted to the psychometric spot in which each subject reached a performance of 80-85%). Stimulus presentation was followed by a black screen for 500 ms. The time-window for participants’ response was 600 ms after target onset. If the participant did not respond within the response window, a screen reading ‘TOO SLOW!’ was presented for 300 ms. Participants were told that if they saw this screen, they should speed up. If a response was made before the deadline, the ‘TOO SLOW!’ screen was omitted and the black screen remained on screen for the 300 ms interval. Finally, each trial ended with presentation of the fixation cross for an additional randomly chosen duration (200, 300 or 400 ms) in order to avoid any habituation or expectation by the subject. Thus, trial duration varied between 1070–1400 ms.

### EEG data acquisition and preprocessing

EEG was recorded with the Starstim system (Neuroelectrics, Cambridge, MA, USA) from 7 positions covering the primary hubs of the fronto-parietal executive control network (Fp1, Fp2, F3, Fz, F4, P3 and P4) with 3.14cm^2^ Ag/AgCl electrodes and digitalized with 24-bit resolution at a sampling frequency of 500 samples/second. EEG data was referenced to the right mastoid. Independent component analysis (ICA) was utilized to identify and remove activity associated with blinks, eye movements, and other artifacts. Data was filtered from 1Hz to 20Hz to remove non-neural physiological activity (skin/sweat potentials) and noise from electrical outlets. Trials were epoched within a time frame of 200ms before and 800ms after the stimulus onset. The mean of the pre-stimulus baseline [-200,0]ms was then subtracted from the entire ERP waveform for each epoch to eliminate any voltage offset.

To create the ERP spectrograms (or ERSP), the Wavelet transform was applied to each singe trial as implemented in EEGlab’s *newtimef* function, with 1 wavelet cycle at the lowest frequency to 10 cycles at the highest, leading to 22 frequency bins logarithmically spaced in the [3, 20]Hz range and 20 linear time bins in the [0, 800]ms range, where 0 represents the onset of the target stimuli in incongruent trials. The input data frames were thus multidimensional arrays of the form [22 Frequency bins] x [20 Time bins] x [7 channels], with 3 minutes of data per subject approximately. For comparison purposes, we also processed with the same parameters a dataset of spontaneous EEG data recorded while the same subjects and ADHD patients were resting with eyes closed (no cognitive task performed).

### Neural network architecture

The DCNN, implemented in Tensorflow^34^, is a relatively simple four layer convolutional network, as shown in Figure 2a. In order to avoid overfitting the data (i.e., overtraining the system to the extent that it negatively impacts the performance of the model on new data), we used the so-called “Dropout” method, a regularization technique in which that randomly selected neurons are ignored during training^35^. The number of iterations in the training process was also limited to the point after which more iterations did not improve training significantly and may lead to overfitting, a method known as “early stopping”^36^. The patch size of the convolutional filter, the pooling parameters and the number of hidden units indicated in Figure 2, as well as the Stochastic Gradient Descent hyper-parameters (number of steps=600, batch size=32), were determined from our previous work using EEG spectrograms^28^, but no fine-tuning or optimization of parameters was applied.

**Figure 2.**
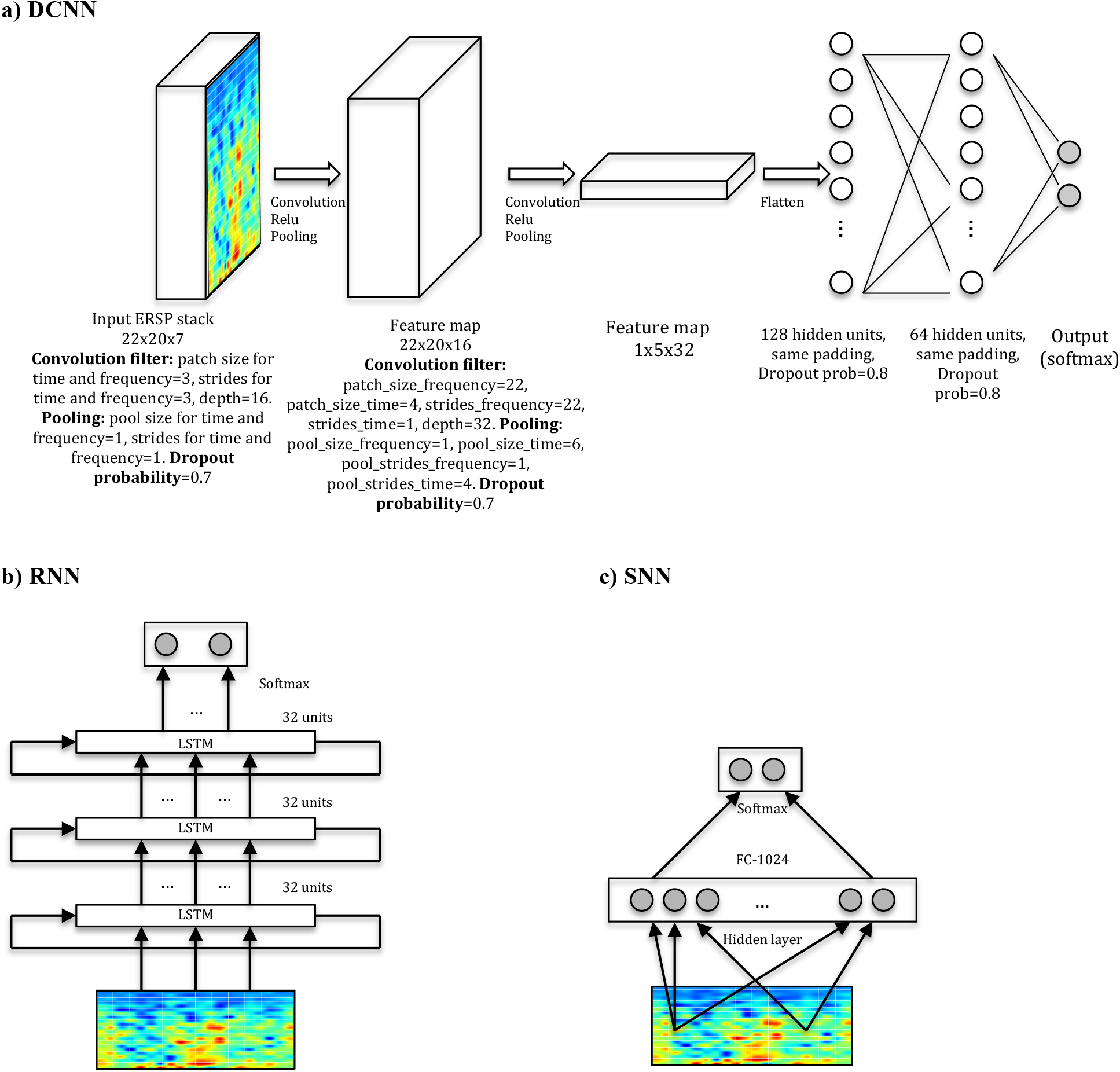
Network architectures. **a) DCNN** model displaying input, convolution with pooling layers, and hidden-unit layers. The first two layers perform the convolution, the Rectified Linear Units (ReLU) function and the pooling processes for feature extraction. The last two layers with 128 and 64 hidden nodes perform the class classification in HC or ADHD. For each trial (or frame), the classifier outputs the probability of the frame belonging to each class (using the softmax function^16^) and, after averaging over frames per subject, we obtained the probability of the subject belonging to each class. Classification was performed by choosing the class with maximal probability. **b) RNN** consisting of three stacked layers of LSTM cells, where each cell uses as input the outputs of the previous one. Each cell used 32 hidden units, and dropout was used to regularize it. **c) SNN** architecture used for comparison with one layer of 1024 units.

We compared the DCNN’s performance with a Shallow Neural Network (SNN) (Figure 2c), a more basic machine neural network with a hidden layer, and with a RNN consisting of stacked LSTM^16,37^, a type of RNN capable of using information about events in the past (memory) to inform predictions in the future (Figure 2b).

### Performance assessment

The performance metrics assessed for each architecture were accuracy (probability of good a classification) and area under the curve (AUC)^38^. Classification performance was validated using the leave-pair out cross-validation (LPO), a method for model selection and performance assessment of deep learning algorithms that consists of training the network N=20×20=400 times (all possible combinations of pairing 1 HC with 1 ADHD), holding one sample from each group out from the training set at a time, and measuring the performance using the held out pair as a test set^38^.

To account for the significant differences in age between the ADHD and HC groups, we applied the Inverse Probability Weighting (IPW) method^39^, which assigns different weights to the subjects in the training process according to their propensity score^40^. The IPW method lead to the same performance without adjustment in all architectures, thus ruling out the effect of age as a confounding factor.

### Feature visualization

Once the network was trained, it was used to find out what type of inputs optimally excite the output nodes using a method popularly known as “DeepDream”, which refers to the generation of synthetic images that produce desired activations in a trained deep network by exaggerating small features within them^30^. The algorithm maximizes a particular class score using gradient descent, starting from a null or random noise image. In particular, we computed the DeepDream spectrograms averaged over N=400 experiments by maximizing the output logits after 30 iterations in steps of 1, initializing with different random images (seeds).

## Results

The results from classification using different methods and datasets are detailed in Figure 3a and 3b, showing that the DCNN trained with ERSPs reached an accuracy of 88% (AUC=96%), very similar to the RNN performance (Accuracy=86%, AUC=95%) and outperforming the SNN (Accuracy=78%, AUC=92%). In comparison with spontaneous EEG spectrograms, ERSPs provided better performance for all architectures. To assess the performance of each individual channel, we also trained the DCNN with ERSP data from single channels and found that frontal (F3, Fz and F4) and parietal electrodes (P3, P4) provide the best performance compared to frontopolar (Fp1, Fp2) electrodes (Figure 3c and 3d).

**Figure 3.**
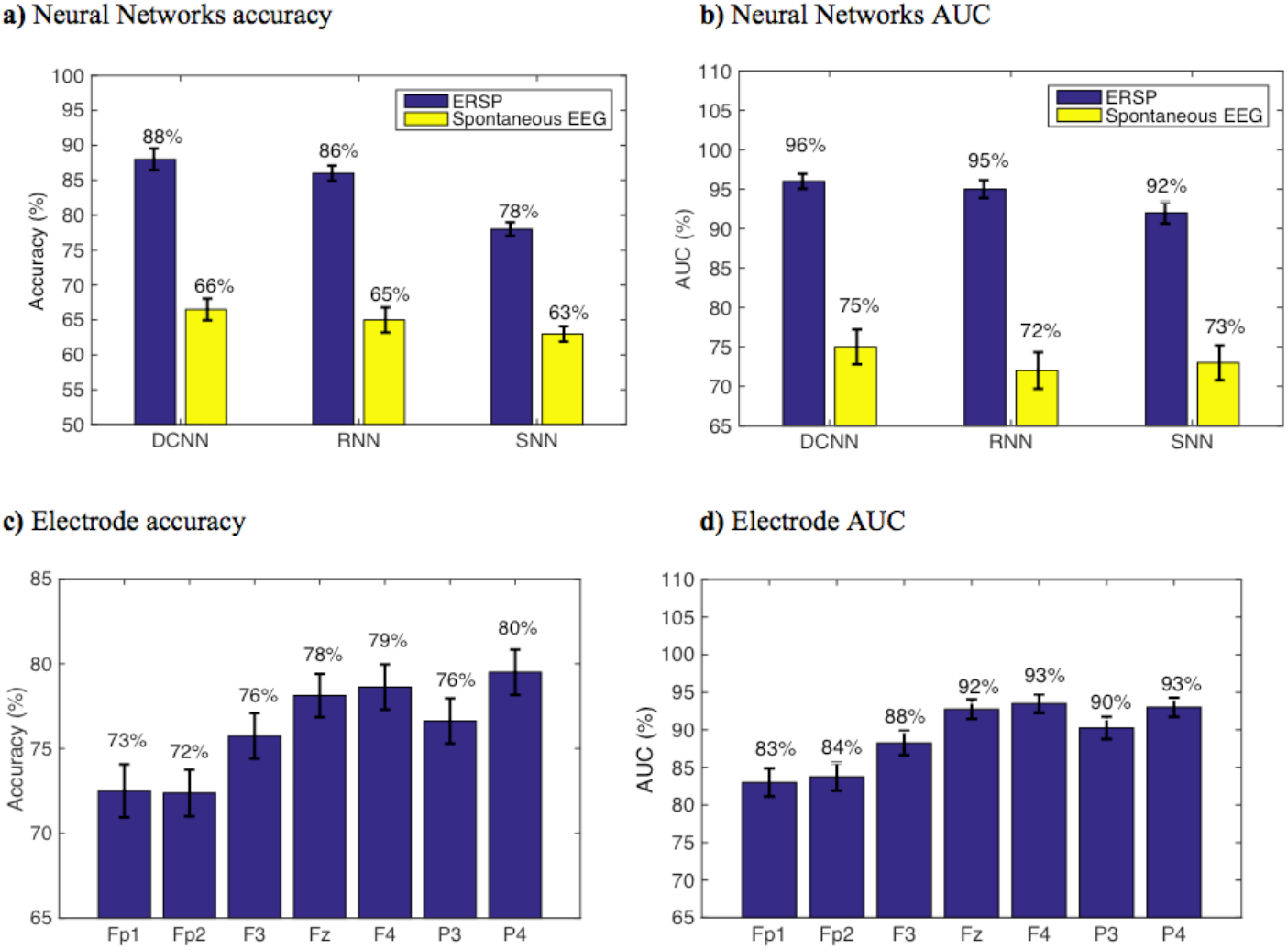
Performance assessment. Neural networks accuracy (a) and AUC (b) with ERSP and spontaneous EEG data. Electrode accuracy (c) and AUC (d) in a DCNN trained with ERSP data from single channels. Error bars indicate mean square error.

The mean DeepDream ERSP averaged over channels can be seen in Figure 4 (see Table S1 for individual channels). The difference between groups reveals that the main feature that optimally excites the network nodes is an increased power for the ADHD group in the delta-theta band (3-7 Hz) around 100 ms and a decreased power in the alpha band (7-12 Hz) along the entire time course, with a residual decrease in theta and beta. Note that the patterns shown in the DeepDream ERSP are very similar to the patterns of the ERSP computed from the real data (Table S1), thus showing that the network is actually learning real neurophysiologically identifiable differences between groups.

**Figure 4.**
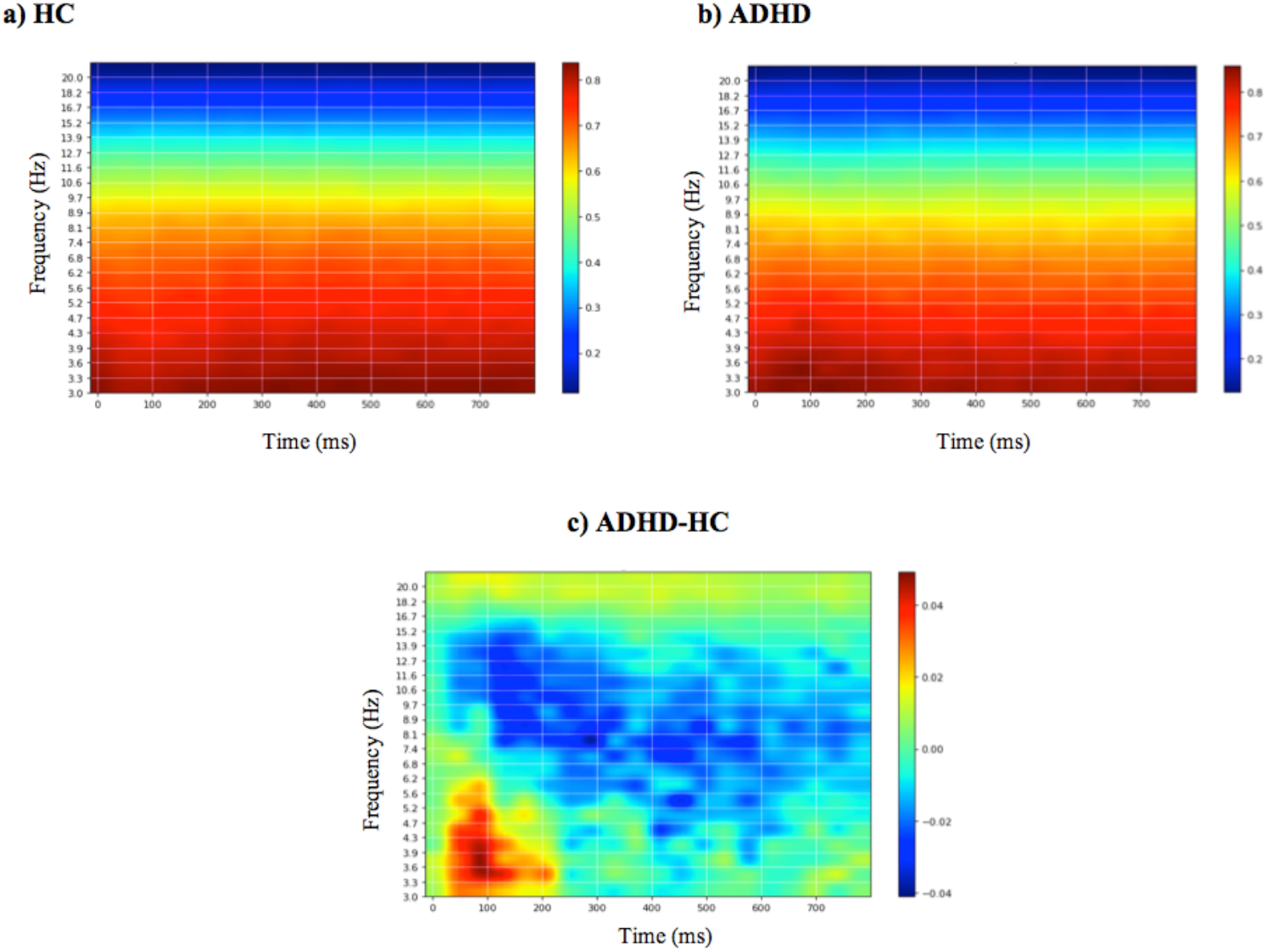
Mean DeepDream ERSP. Mean DeepDream ERSP averaged over channels generated after 400 experiments for healthy controls (a), ADHD patients (b) and their difference (c). Color bar units = dB.

Behaviorally, the mean reaction time was significantly slower for ADHD compared to HC (RT_ADHD_=368ms, RT_HC_=321ms, β=46ms, CI=[38,53]ms, p<0.001), which can be expected with this type of population with attention deficits, but the mean percentage of responses for each group was not significantly different (Accuracy_ADHD_=62%, Accuracy_HC_=65%, β=0.12, CI=[0.02, 0.26], p=0.10).

## Discussion

In this study we present a viable deep learning model for effective discrimination of patients with ADHD, providing a new tool for the analysis of EEG dynamics in ADHD and supporting the potential of deep learning strategies for biomarker development in neuropsychiatry. We deem this approach to be particularly interesting for various reasons. First, it largely mitigates the need for EEG feature selection (spectral bands, time ranges, specific ERP components and channels). Second, results with ERSPs represent an improvement over spontaneous EEG spectrograms (e.g. subject Accuracy with DCNN was 88% for ERSP vs. 66% for spontaneous resting-state data). Third, the performances of the proposed DCNN and RNN systems are very similar and they outperform the SNN used for comparison. Finally, through the use of feature visualization, we identify neurophysiologically interpretable features that can be extracted from the model, providing further validation and evidence that the network performance is not driven by noise or artifact signals in the data and providing a mechanistic model with added value to understand pathophysiology.

The higher accuracy provided by DCNN and RNN compared to SNN proves that the complex deep approaches with more layers and units provide better performance than more shallow networks. The similar performance of DCNN compared to RNN shows, however, that the higher computational demands of RNN do not provide better performance than the DCNN approaches, thus proving DCNN as a more efficient method than RNN.

The fact that ERSP data provide better performance than spontaneous EEG data with all architectures also shows that event-related data from a highly yield task that elicits the primary executive functions impaired in ADHD is a better predictor than spontaneous EEG data recorded while the participants are at resting state.

Finally, through the use of feature visualization we show that the main spectral features picked up by the DCNN nodes are a decrease in alpha activity over the entire time course and an increased delta-theta activity around 100 ms for ADHD patients compared to HC. There is evidence that an increased alpha activity (or alpha Event-Related Synchronization, ERS) in conflict and inhibitory tasks is related with an improved inhibition of the prepotent response, reflecting a top-down inhibitory control process^41^. Therefore, we interpret the decrease in alpha power in ADHD as a deficit in cognitive control. On the other hand, the increased delta-theta activity is localized to 100ms and is probably related to the increase in N100 amplitude in the time domain (Table S1). N100 is a visual sensory evoked potential that is thought to index sensory analysis of simple stimulus features and whose amplitude is influenced by selective attention^42^. The increased delta-theta power in that latency suggests that ADHD patients manifest specific alterations in the process of early selection of visual task stimuli^43^. Given that there were no significant differences in the percentage of correct responses between ADHD and HC, we interpret this as a compensation strategy to offset inhibitory deficits by shifting more attention to the task^44,45^.

Note that the DeepDream spectrograms generated for Fp1 and Fp2 are substantially different and provide lower performance that the other positions (F3, F4, Fz, P3, P4), which may be explained by the lower signal quality of frontopolar positions due to blinks, muscle artefacts and sweat. The lower performance of F3 and P3 electrodes compared to F4 and P4 may also be related to the lower power scale in their DeepDream spectrogram, respectively (Table S1). This may suggest the existence of inter-hemispheric differences in the features driving the discrimination between ADHD and HC.

Similar studies have explored the application of deep learning to EEG signals. For example, DCNNs have been used for epilepsy prediction and monitoring46, mental workload classification47 and motor imagery classification48-50. Deep neural networks have also shown convincing results in classifying psychiatric disorders such as dementia^51^ and ADHD^52-57^, mostly with MRI data. To our knowledge, this is the first study using a deep learning approach with EEG event-related spectral data to discriminate adult ADHD patients from HC with no prior selection of EEG features and the combination with feature visualization techniques to provide further mechanistic evidence of the underlying pathophysiology driving the classification. This is particularly important, as it not only allows to develop clinical tools but also to delineate pathological signatures and disease mechanisms.

One of the limitations of this study is the relatively small size of the dataset, with the consequent limitation on the network due to susceptibility to overfitting. Although “early stopping” and regularization should mitigate this issue, further improvements could be achieved with bigger datasets. Another limitation is the age difference between the two groups. While the mean ages are well after the period of brain maturation when myelination and ADHD symptoms are still changing, and well before a geriatric threshold when other type of biological changes (including normal aging) may affect cognition, we addressed this possible confounder using Inverse Probability Weighting. The age difference was an artifact caused by the fact that the two cohorts were recruited prospectively for independent studies (though at the same time and with the same exact protocol and hardware) and then analyzed together retrospectively to address the proposed questions, hence the lack of appropriately age-matched controls. Future prospective validation studies should use larger cohorts and randomize age-matched controls.

It is also worth mentioning that, although the current work considerably eliminates the need for manual extraction of features, it is still focused on classification during high yield incongruent trials of a specific task. While this requires a priori knowledge constrains, if validated with higher definition EEG and bigger datasets, it may be a helpful diagnostic and biomarker development strategy (i.e. choosing high yield events of a high yield task) with practical future procedural advantages (i.e. it would be easy to implement it in clinical settings with currently existing tools, such as tasks for neuropsychological assessments and standard EEG for electrophysiological diagnosis).

Our findings may also have several implications from the clinical perspective by bringing new information to inform the clinician’s decisions. Although the networks in this study have been trained with a small dataset of 40 subjects, if validated with bigger datasets this approach could be used to support the diagnosis of ADHD on a single-patient basis. The fact that the current networks have been trained with low-resolution EEG datasets (7 channels) of short duration (3 minutes) would make it easy to implement them not only in an EEG clinical unit, but possibly by an outpatient clinician, eliminating the need to get longer or higher quality data with sophisticated and clinically unpractical EEG systems. However, even if these deep learning systems are properly validated in the future, clinicians should view their output as statistical predictions, not as a ground truth, and they should judge whether the prediction applies to that specific patient and decide if additional data or expertise is needed to inform that decision.

Future work should include the exploration of this approach with larger datasets as well as a more systematic study of network architecture and regularization schemes. This includes the use of deeper architectures, improved data augmentation methods, alternative data segmentation and normalization schemes. With regards to data preprocessing, we should consider improved spectral estimation using more advanced techniques such as state-space estimation and multitapering^58^, and the use of cortical or scalp-mapped EEG data prior creation of spectrograms.

Finally, we note that we make no attempt to fully-optimize our architecture in this study. In particular, no fine-tuning of hyper-parameters has been carried out using a validation set approach, a task we reserve for future work with larger datasets. Our aim was to validate the idea that deep learning approaches can provide value for the analysis of time-frequency representations of EEG, and particularly ERSP data, for the effective discrimination of ADHD.

## Data Availability

The datasets generated and analyzed during the current study are available from the senior author (JAC) on reasonable request.

## Acknowledgements

This research was partly supported by NIH grants (RO1 MH112737, R21 DA042271, R21 AG056958 and R21 MH113018) and the Louis V. Gerstner III Research Scholar Award to JAC.

## Author contributions

LD contributed with the processing of the data, the implementation of the deep learning systems, and the writing of the article. GR contributed with the conception and design of the deep learning systems and the revision of the manuscript. JC contributed with the conception and design of the study, the supervision of data acquisition and the findings, and the critical revision of the manuscript. All authors gave the approval to the final version of the manuscript to be published.

## References

1 Shamay-Tsoory, S. G. & Aharon-Peretz, J. Dissociable prefrontal networks for cognitive and affective theory of mind: a lesion study. Neuropsychologia 45, 3054–3067, doi:10.1016/j.neuropsychologia.2007.05.021 (2007).

2 Biederman, J. et al. Functional impairments in adults with self-reports of diagnosed ADHD: A controlled study of 1001 adults in the community. The Journal of clinical psychiatry 67, 524–540 (2006).

3 Fayyad, J. et al. Cross-national prevalence and correlates of adult attention-deficit hyperactivity disorder. The British journal of psychiatry : the journal of mental science 190, 402–409, doi:10.1192/bjp.bp.106.034389 (2007).

4 Kessler, R. C. et al. The prevalence and correlates of adult ADHD in the United States: results from the National Comorbidity Survey Replication. The American journal of psychiatry 163, 716–723, doi:10.1176/ajp.2006.163.4.716 (2006).

5 Polanczyk, G., de Lima, M. S., Horta, B. L., Biederman, J. & Rohde, L. A. The worldwide prevalence of ADHD: a systematic review and metaregression analysis. The American journal of psychiatry 164, 942–948, doi:10.1176/ajp.2007.164.6.942 (2007).

6 Jasper, H. H., Solomon, P. & Bradley, C. Electroencephalographic analyses of behavior problem children. American Journal of Psychiatry 95, 641–658, doi:10.1176/ajp.95.3.641 (1938).

7 Lenartowicz, A. & Loo, S. K. Use of EEG to Diagnose ADHD. Current psychiatry reports 16, 498, doi:10.1007/s11920-014-0498-0 (2014).

8 Fabiani, M., Gratton, G. & Federmeier, K. D. in Handbook of psychophysiology, 3rd ed. 85–119 (Cambridge University Press, 2007).

9 Sanei, S. & Chambers, J. A. EEG Signal Processing. (John Wiley & Sons Ltd, 2013).

10 Woodman, G. F. A brief introduction to the use of event-related potentials in studies of perception and attention. Attention, perception & psychophysics 72, 2031–2046, doi:10.3758/app.72.8.2031 (2010).

11 Pokryszko-Dragan, A. et al. Event-related potentials and cognitive performance in multiple sclerosis patients with fatigue. Neurological sciences : official journal of the Italian Neurological Society and of the Italian Society of Clinical Neurophysiology 37, 1545–1556, doi:10.1007/s10072-016-2622-x (2016).

12 Paulraj, M. P., Subramaniam, K., Yaccob, S. B., Adom, A. H. & Hema, C. R. Auditory evoked potential response and hearing loss: a review. The open biomedical engineering journal 9, 17–24, doi:10.2174/1874120701509010017 (2015).

13 Durstewitz, D., Koppe, G. & Meyer-Lindenberg, A. Deep neural networks in psychiatry. Molecular Psychiatry, doi:10.1038/s41380-019-0365-9 (2019).

14 Kroupi, E. et al. in HBP Student Conference - Transdisciplinary Research Linking Neuroscience, Brain Medicine and Computer Science (Viena, Austria, 2017).

15 Ruffini, G., Ibañez, D., Castellano, M., Dunne, S. & Soria-Frisch, A. EEG-driven RNN Classification for Prognosis of Neurodegeneration in At-Risk Patients. Artificial Neural Networks and Machine Learning – ICANN 2016, 306-313 (2016).

16 Goodfellow, I., Bengio, Y. & Courville, A. Deep Learning. (MIT Press, 2016).

17 Mueller, A., Candrian, G., Kropotov, J. D., Ponomarev, V. A. & Baschera, G. M. Classification of ADHD patients on the basis of independent ERP components using a machine learning system. Nonlinear Biomedical Physics 4, S1, doi:10.1186/1753-4631-4-s1-s1 (2010).

18 Tenev, A. et al. Machine learning approach for classification of ADHD adults. International journal of psychophysiology : official journal of the International Organization of Psychophysiology 93, 162–166, doi:10.1016/j.ijpsycho.2013.01.008 (2014).

19 Jahanshahloo, H. R., Shamsi, M., Ghasemi, E. & Kouhi, A. Automated and ERP-Based Diagnosis of Attention-Deficit Hyperactivity Disorder in Children. Journal of Medical Signals and Sensors 7, 26–32 (2017).

20 Nazhvani, A. D., Boostani, R., Afrasiabi, S. & Sadatnezhad, K. Classification of ADHD and BMD patients using visual evoked potential. Clinical neurology and neurosurgery 115, 2329–2335, doi:10.1016/j.clineuro.2013.08.009 (2013).

21 Sadatnezhad, K., Boostani, R. & Ghanizadeh, A. Classification of BMD and ADHD patients using their EEG signals. Expert Systems with Applications 38, 1956–1963, doi: https://doi.org/10.1016/j.eswa.2010.07.128 (2011).

22 Ahmadlou, M. & Adeli, H. Wavelet-synchronization methodology: a new approach for EEG-based diagnosis of ADHD. Clinical EEG and neuroscience 41, 1–10, doi:10.1177/155005941004100103 (2010).

23 Abibullaev, B. & An, J. Decision support algorithm for diagnosis of ADHD using electroencephalograms. Journal of medical systems 36, 2675–2688, doi:10.1007/s10916-011-9742-x (2012).

24 Ruffini, G. et al. Algorithmic complexity of EEG for prognosis of neurodegeneration in idiopathic rapid eye movement behavior disorder (RBD). bioRxiv (2018).

25 Oord, A. v. d., Dieleman, S. & Schrauwen, B. in Proceedings of the 26th International Conference on Neural Information Processing Systems - Volume 2 2643–2651 (Curran Associates Inc., Lake Tahoe, Nevada, 2013).

26 Tsinalis, O., M. Matthews, P., Guo, Y. & Zafeiriou, S. Automatic Sleep Stage Scoring with Single-Channel EEG Using Convolutional Neural Networks. (2016).

27 Vilamala, A., Madsen, K. H. & Hansen, L. K. Deep Convolutional Neural Networks for Interpretable Analysis of EEG Sleep Stage Scoring. 2017 International workshop on Machine Learning for signal processing (2017).

28 Ruffini, G. et al. Deep Learning With EEG Spectrograms in Rapid Eye Movement Behavior Disorder. Frontiers in neurology 10, 806 (2019).

29 Dauwels, J., Vialatte, F., Musha, T. & Cichocki, A. A comparative study of synchrony measures for the early diagnosis of Alzheimer’s disease based on EEG. NeuroImage 49, 668–693, doi: https://doi.org/10.1016/j.neuroimage.2009.06.056 (2010).

30 Alexander, M., Christopher, O. & Mike, T. Inceptionism: Going Deeper into Neural Networks. Google Research Blog (2015).

31 American-Psychiatric-Association. The Diagnostic and Statistical Manual of Mental Disorders. 5th Edition edn, (American Psychiatric Publishing, 2013).

32 Kessler, R. C. et al. The World Health Organization Adult ADHD Self-Report Scale (ASRS): a short screening scale for use in the general population. Psychol Med 35, 245–256 (2005).

33 Kopp, B., Rist, F. & Mattler, U. N200 in the Flanker task as a neurobehavioral tool for investigating executive control. Psychophysiology 33, 282–294 (1996).

34 Abadi, M. et al. in Proceedings of the 12th USENIX conference on Operating Systems Design and Implementation 265-283 (USENIX Association, Savannah, GA, USA, 2016).

35 Srivastava, N., Hinton, G., Krizhevsky, A., Sutskever, I. & Salakhutdinov, R. Dropout: A Simple Way to Prevent Neural Networks from Overfitting. Journal of Machine Learning Research 15, 1929–1958 (2014).

36 Prechelt, L. in Neural Networks: Tricks of the Trade (eds Genevieve B. Orr & Klaus-Robert Müller) 55–69 (Springer Berlin Heidelberg, 1998).

37 Hochreiter, S. & Schmidhuber, J. Long Short-Term Memory. Neural Computation 9, 1735–1780, doi:10.1162/neco.1997.9.8.1735 (1997).

38 Antti, A., Tapio, P., Willem, W., Bernard De, B. & Tapio, S. 3–13 (PMLR, 2009).

39 Linn, K. A., Gaonkar, B., Doshi, J., Davatzikos, C. & Shinohara, R. T. Addressing Confounding in Predictive Models with an Application to Neuroimaging. The international journal of biostatistics 12, 31–44, doi:10.1515/ijb-2015-0030 (2016).

40 Austin, P. C. An Introduction to Propensity Score Methods for Reducing the Effects of Confounding in Observational Studies. Multivariate behavioral research 46, 399–424, doi:10.1080/00273171.2011.568786 (2011).

41 Klimesch, W., Sauseng, P. & Hanslmayr, S. EEG alpha oscillations: The inhibition–timing hypothesis. Brain Research Reviews 53, 63–88, doi: https://doi.org/10.1016/j.brainresrev.2006.06.003 (2007).

42 Rugg, M. D., Milner, A. D., Lines, C. R. & Phalp, R. Modulation of visual event-related potentials by spatial and non-spatial visual selective attention. Neuropsychologia 25, 85–96 (1987).

43 Yordanova, J., Heinrich, H., Kolev, V. & Rothenberger, A. Increased event-related theta activity as a psychophysiological marker of comorbidity in children with tics and attention-deficit/hyperactivity disorders. NeuroImage 32, 940–955, doi: https://doi.org/10.1016/j.neuroimage.2006.03.056 (2006).

44 Broyd, S. J. et al. The effect of methylphenidate on response inhibition and the event-related potential of children with attention deficit/hyperactivity disorder. International journal of psychophysiology : official journal of the International Organization of Psychophysiology 58, 47–58, doi:10.1016/j.ijpsycho.2005.03.008 (2005).

45 Prox, V., Dietrich, D. E., Zhang, Y., Emrich, H. M. & Ohlmeier, M. D. Attentional processing in adults with ADHD as reflected by event-related potentials. Neuroscience Letters 419, 236–241 (2007).

46 Liang, J., Lu, R., Zhang, C. & Wang, F. in IEEE International Conference on Healthcare Informatics (ICHI) 184-191 (2016).

47 Ma, T. et al. The extraction of motion-onset VEP BCI features based on deep learning and compressed sensing. J Neurosci Methods 275, 80–92, doi:10.1016/j.jneumeth.2016.11.002 (2017).

48 Bashivan, P., Rish, I., Yeasin, M. & Codella, N. Learning Representations from EEG with Deep Recurrent-Convolutional Neural Networks. (2015).

49 Tabar, Y. R. & Halici, U. A novel deep learning approach for classification of EEG motor imagery signals. J Neural Eng 14, 016003, doi:10.1088/1741-2560/14/1/016003 (2017).

50 An, X., Kuang, D., Guo, X., Zhao, Y. & He, L. in Intelligent Computing in Bioinformatics. (eds De-Shuang Huang, Kyungsook Han, & Michael Gromiha) 203–210 (Springer International Publishing).

51 Vieira, S., Pinaya, W. H. & Mechelli, A. Using deep learning to investigate the neuroimaging correlates of psychiatric and neurological disorders: Methods and applications. Neurosci Biobehav Rev 74, 58–75, doi:10.1016/j.neubiorev.2017.01.002 (2017).

52 Deshpande, G., Wang, P., Rangaprakash, D. & Wilamowski, B. Fully Connected Cascade Artificial Neural Network Architecture for Attention Deficit Hyperactivity Disorder Classification From Functional Magnetic Resonance Imaging Data. IEEE transactions on cybernetics 45, 2668–2679, doi:10.1109/tcyb.2014.2379621 (2015).

53 Han, X., Zhong, Y., He, L., Yu, P. S. & Zhang, L. in Brain Informatics and Health. (eds Yike Guo et al.) 156–166 (Springer International Publishing).

54 Hao, A. J., He, B. L. & Yin, C. H. in 2015 IET International Conference on Biomedical Image and Signal Processing (ICBISP 2015). 1–6.

55 Kuang, D. & He, L. in 2014 International Conference on Cloud Computing and Big Data. 27–32.

56 Kuang, D., Guo, X., An, X., Zhao, Y. & He, L. in Intelligent Computing in Bioinformatics. (eds De-Shuang Huang, Kyungsook Han, & Michael Gromiha) 225-232 (Springer International Publishing).

57 Zou, L., Zheng, J., Miao, C., Mckeown, M. J. & Wang, Z. J. 3D CNN Based Automatic Diagnosis of Attention Deficit Hyperactivity Disorder Using Functional and Structural MRI. IEEE Access 5, 23626–23636, doi:10.1109/ACCESS.2017.2762703 (2017).

58 Kim, S.-E., Behr, M. K., Ba, D. & Brown, E. N. State-space multitaper time-frequency analysis. Proceedings of the National Academy of Sciences 115, E5, doi:10.1073/pnas.1702877115 (2018).

